# Generalizable Deep Learning Framework for Radiotherapy Dose Prediction Across Cancer Sites, Prescriptions and Treatment Modalities

**DOI:** 10.64898/2026.04.17.26350770

**Authors:** Ho-hsin Chang, Rex Alexander Cardan, Ritish Nedunoori, John B. Fiveash, Richard A. Popple, Sandeep Bodduluri, Dennis Stanley, Joseph Harms, Carlos E. Cardenas

## Abstract

Optimizing radiotherapy dose distributions remain a resource-intensive bottleneck. Existing AI-based dose prediction methods often have limited generalizability because they rely on small, heterogeneous datasets. We present nnDoseNetv2, an auto-configured, end-to-end framework for dose prediction across diverse disease sites (head and neck, prostate, breast, and lung), prescription levels (1.5–84 Gy), and treatment modalities (IMRT, VMAT, and 3D-CRT). By integrating machine-specific beam geometry with 3D structural information, the framework is designed to generalize across varied clinical scenarios.

A single multi-site model was trained on 1,000 clinical plans. On sites seen during training, performance was comparable to specialized site-specific models. On unseen sites (liver and whole brain), the model outperformed site-specific models, with mean absolute errors of 2.46% and 6.97% of prescription, respectively.

These results suggest that geometric awareness can bridge disparate anatomical domains while eliminating the need for site-specific model maintenance, providing a scalable and high-fidelity approach for personalized radiotherapy planning.

## 1 Introduction

Radiotherapy (RT) treatment planning is a complex, iterative process that can take days to weeks. Planners begin by configuring the treatment planning system (TPS) with beam parameters (e.g. delivery machine, number of beams, beam angles, etc.) which strongly influence the final dose distribution. To reach a desirable plan, planners iteratively run TPS optimizations and adjust the planning goal to balance the trade-off between tumor coverage and sparing healthy tissue. Due to time and resource constraints, planners routinely face a compromise between plan quality and turnaround time. Therefore, a fast, automated deep learning (DL) dose prediction tool could reduce planning time from weeks to minutes by providing dose estimation and potential clinical goal for the RT planning.

The use of DL tools to assist RT workflows is becoming increasingly accepted ^1–3^. However, DL research for dose prediction has been dominated by site-specific models^4–29^. These models are typically trained on homogeneous datasets with minimal variation in delivery machines, disease sites, or prescriptions. Such a fragmented data selection strategy increases the technical burden of model management; maintaining a distinct model for every clinical scenario is unsustainable for most centers, particularly for resource-limited clinics with constrained hardware and data access. Furthermore, this fragmentation hinders the generalizability and robustness of trained models by limiting both data diversity and the total scale of training data. Currently, the largest public dataset remains the 2020 OpenKBP challenge dataset^30,31^, which comprises 340 processed head and neck 9-field intensity modulated radiation therapy (IMRT) plans containing a primary 70 Gy target with optional targets of 63 Gy and 56 Gy. Therefore, a beam aware, multi-site training approach utilizing heterogeneous data is crucial to improve generalizability, robustness and scale of the model and further improve the advancing global oncology care.

Our previous work, nnDoseNet, addressed the technical barriers of DL implementation by providing an off-the-shelf, auto-configuring framework. By automatically tailoring preprocessing and training pipelines to specific data distributions and available GPU memory, nnDoseNet enables clinics without high-performance computing (HPC) infrastructure or specialized programming expertise to develop robust models. This approach recognizes that clinical goals and planning strategies vary significantly across institutions; rather than providing a static, pre-trained model, this flexible framework allows clinics to train tailored models using their own data. By providing an end-to-end training pipeline that processes data directly from the DICOM format, the barrier to entry for AI adoption is significantly lowered. However, while nnDoseNet streamlined model management through this systemic pipeline, it was primarily developed using homogeneous datasets. It lacked the capacity to handle multi-prescription, cross-site, or multi-modality scenarios because the original framework could not effectively extract and incorporate beam geometry from the plan DICOM.

While recent studies have incorporated beam geometry to validate cross-site performance, these approaches often lack publicly available code or a standardized, end-to-end framework. This lack of open-resource tools prevents other institutions from adopting these tools.

With the nnDoseNet^5^ pipeline and a sufficiently large dataset, we hypothesize that a single model that handles multiple delivery machines, disease sites, and prescriptions, is feasible. In this research, a multi-site model is trained with a large-scale, multi-site dataset (HN, prostate, breast, and lung) consisting of 1,000 clinically delivered RT plans at our institution, making this the first deep learning dose predictor trained on a multi-site dataset of this scale. The multi-site model, trained with nnDoseNet, and tested on an independent set of 300 cases from seen and unseen disease site. Our results show that the multi-site model achieves comparable (and better in some cases) accuracy than site-specific models, demonstrating the feasibility of a general model without loss of performance. To our knowledge, this is the first demonstration of a deep learning model trained across such diverse RT planning scenarios.

## 2 Materials and Methods

### 2.1 Large Scale Multi-Site Dataset

An institutional dataset comprising 1,300 cases across six anatomical sites was utilized for this study, with all records anonymized via DICOMAnon™ Ultimate (Red Ion LLC). Each case includes a comprehensive suite of RT records in DICOM format, including planning CT images, structure sets (organs-at-risk and targets), clinically delivered treatment plans, and corresponding 3D dose distributions. The dataset reflects real-world clinical heterogeneity, consisting of a mixture of 3D conformal radiotherapy (3D-CRT), IMRT, and volumetric modulated arc therapy (VMAT).

To evaluate the model’s performance across diverse clinical scenarios, we incorporated four seen disease sites (head and neck [HN], prostate, breast, and lung) for model training and initial testing, alongside two unseen sites (liver and whole brain) reserved exclusively for external validation. The training cohort consisted of 1,000 cases (250 per seen site), while the testing cohort comprised 300 cases (50 per seen site and 50 per unseen site).

As illustrated in Figure 1, the training and testing subsets for these sites exhibit nearly identical distributions across all six clinical parameters (dose per fraction, total dose, etc). This distributional consistency ensures that the testing set is representative of the training data diversity, providing a reliable measure of model performance. The data complexity is further evidenced by the wide range of targets and prescriptions across sites: the prostate dataset includes 1–4 targets (18–84 Gy), breast includes 1–5 targets (2.5–50 Gy), lung includes 1–4 targets (1.5–66 Gy), and HN includes 1–4 targets (2–70 Gy). Variations in prescription and fractionation reflect diverse clinical intents, spanning boost phases, stereotactic treatments, conventional radiotherapy, and palliative regimens.

**Figure 1.**
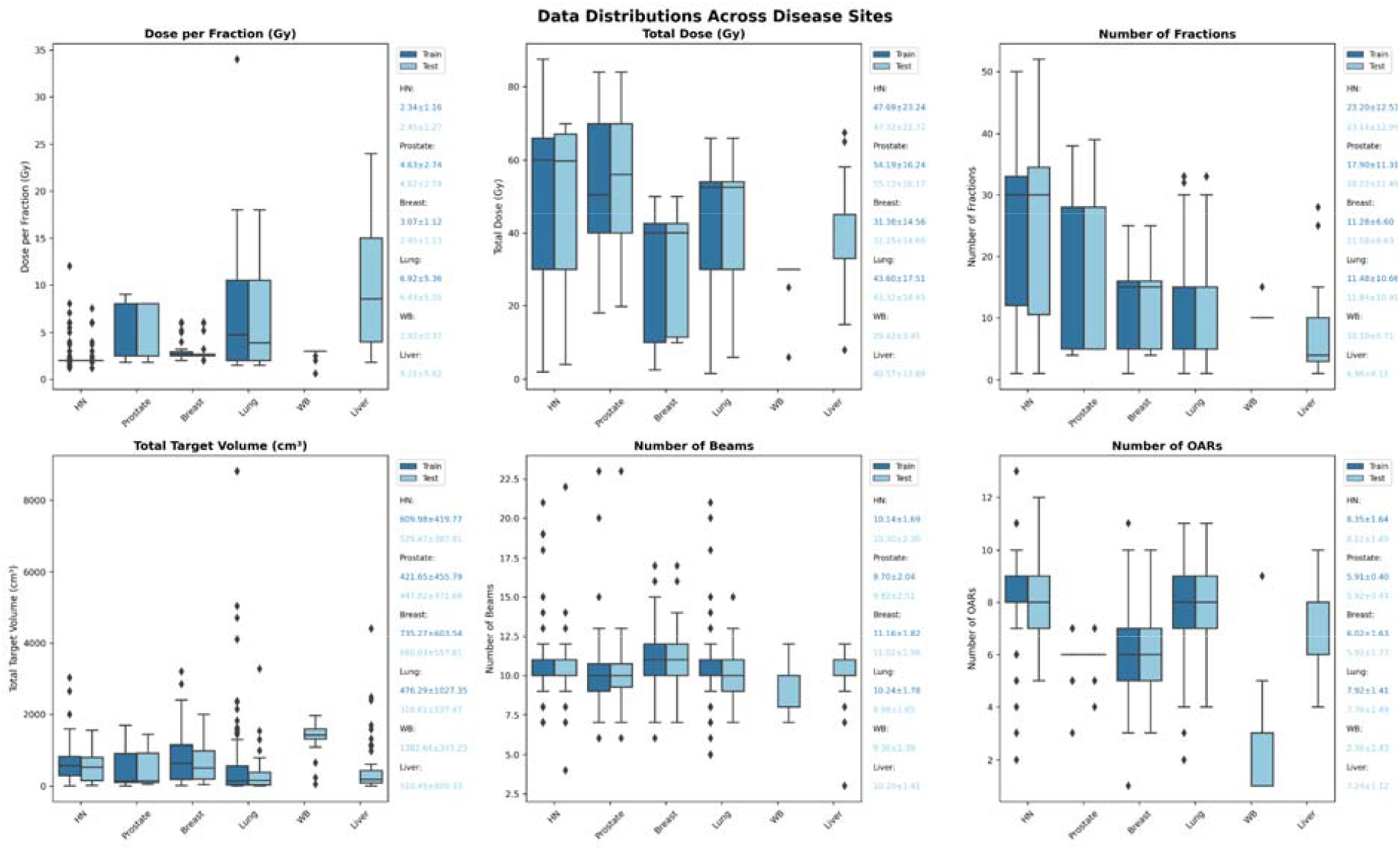
Characterization of dataset diversity across disease sites. Distribution of key radiotherapy parameters for the six disease sites included in this study. The head and neck (HN), prostate, breast, and lung cohorts served as seen sites for both model training and testing. The whole brain (WB) and liver cohorts were reserved as unseen sites for independent validation of model generalizability. The plots illustrate the wide variance in prescription doses, target counts, and treatment modalities, highlighting the heterogeneity required for a universal dose prediction framework. Notably, the training and testing distributions for the four seen sites are closely matched, demonstrating an even data split that maintains representative complexity across both cohorts.

**Figure 2.**
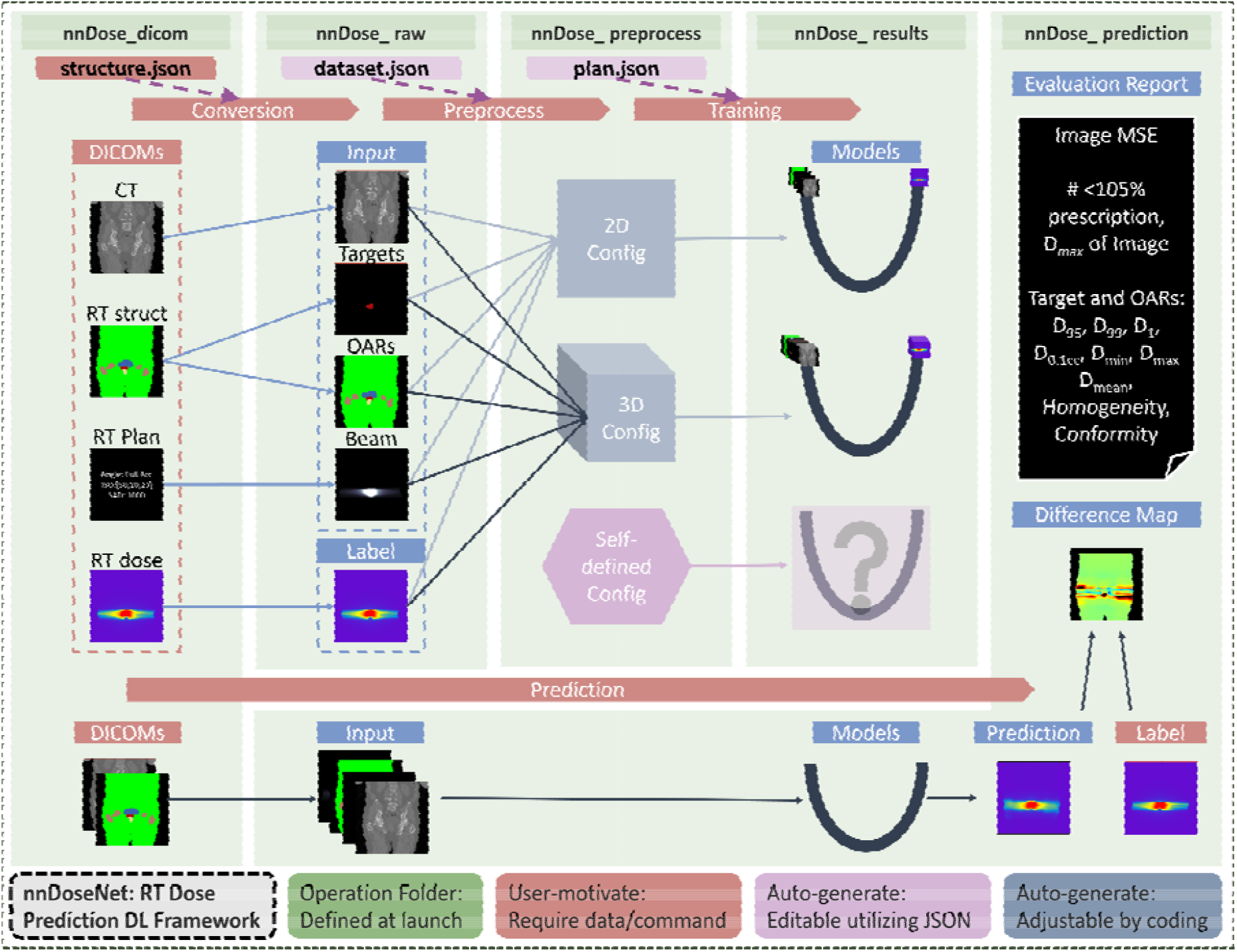
Integrated nnDoseNet framework for automated, beam-aware dose prediction. The upgraded pipeline incorporates a module for extracting machine-specific beam geometry from RTPLAN files and integrating it directly into the data conversion process. nnDoseNet provides an end-to-end architecture that auto-configure data preprocessing, systematic model training, and robust dose evaluation pipeline. The framework simplifies complex DL workflows by reducing model training to a single-command interface and enabling rapid architectural tuning via standardized JSON configuration files.

### 2.2 Data Preparation

#### 2.2.1 CT and RT DOSE DICOM Extraction

The data conversion pipeline is built upon the established nnDoseNet framework, utilizing an automated workflow to translate DICOM radiotherapy files into NIfTI volumes to standardize the spatial metadata for the DL pipeline. CT images are generated from CT DICOM files, and ground-truth dose maps are extracted from RTDOSE files.

Anatomical structures are processed via the RTSTRUCT conversion module, which extracts, encodes, and separates targets and organs-at-risk (OARs) into corresponding input channels. For this project, the pipeline was configured to recognize 18 key anatomical structures across different disease sites: the body, spine, lungs, esophagus, heart, liver, bladder, rectum, femoral heads, bowel, thyroid, trachea, carina, main bronchi, mandible, brainstem, larynx, and parotids.

For target and dose encoding, the framework employs a two labeling strategy in target designed to evaluate sensitivity to absolute magnitude versus relative distribution:

1. Prescription-labeled (cGy): Target structures are encoded by assigning specific prescription values directly to each constituent voxel (e.g., 70 Gy is encoded as 7,000 cGy), with ground-truth dose maps maintained in absolute Gy.
2. Percentage-labeled (%): Both target masks and ground-truth dose maps are normalized to the maximum target prescription (0 to 100%). This approach is specifically designed to eliminate training bias toward high-dose prescriptions.

#### 2.2.2 RTPLAN and Beam Geometry

The major difference between nnDoseNetv1 and nnDoseNetv2, a beam geometry extraction pipeline leveraging RTPLAN DICOM files is being utilized. By analyzing the isocenter coordinates, source-axis distance (SAD), and beam angles, each beam’s geometry was projected onto the CT volume and created a 3D beam geometry map or a distance-to-beam axis representation. For cross-section area shape, the field shape for beam projection is utilized in 3D-CRT plans and beams-eye-view (BEV) is utilized in IMRT and VMAT. This beam geometry map encodes the machine-specific beam arrangement and is used as an additional input compared to the previous version of nnDoseNet. This beam geometry extraction will be integrated into nnDoseNet as part of future work.

#### 2.2.3 Efficiency Improvement

To enhance computational throughput and reduce memory overhead, several architectural optimizations were implemented. First, the body contour—previously handled as an isolated channel—was consolidated directly into the OAR map, reducing the overall input dimensionality to a streamlined four-channel architecture. Second, all images were resampled to match the native resolution of the dose maps, avoiding the overhead of high-resolution CT grids. Finally, volumes were cropped to a region of interest (ROI) extending ±5 slices cranio-caudally beyond the target boundaries, focusing the model’s learning capacity on the primary irradiated regions while excluding distant anatomical structures that do not contribute to the dose distribution.

#### 2.2.4 Auto-configured Image Preprocess Pipeline

The same intensity preprocessing as nnU-Net^32^ were applied, that the CT were clipped to the [5th, 95th] percentile range of foreground image intensities. The beam geometry map was normalized values to [0,1]. The target and OAR channels were used as label channels (i.e. integers, no intensity normalization applied). For dose maps, the data can be pre-processed in two formats: 1) dose values in cGy (prescription-labeled), and 2) normalized dose values by highest target prescription (percentage-labeled).

### 2.3 Training

#### 2.3.1 Experiments of Training Single-site and Multi-site Models

Two experiments were conducted to train our model: a single-site experiment and a multi-site experiment (Figure 3). In the single-site experiment, four individual models were trained/validated on 250 plans from one site and tested on 50 held-out cases from that same site (total of 200 test cases across four training sites). In the multi-site experiment, models were trained/validated on 1000 plans (250 from each site combined) and tested it on the same 200 held-out cases (50 per site).

**Figure 3.**
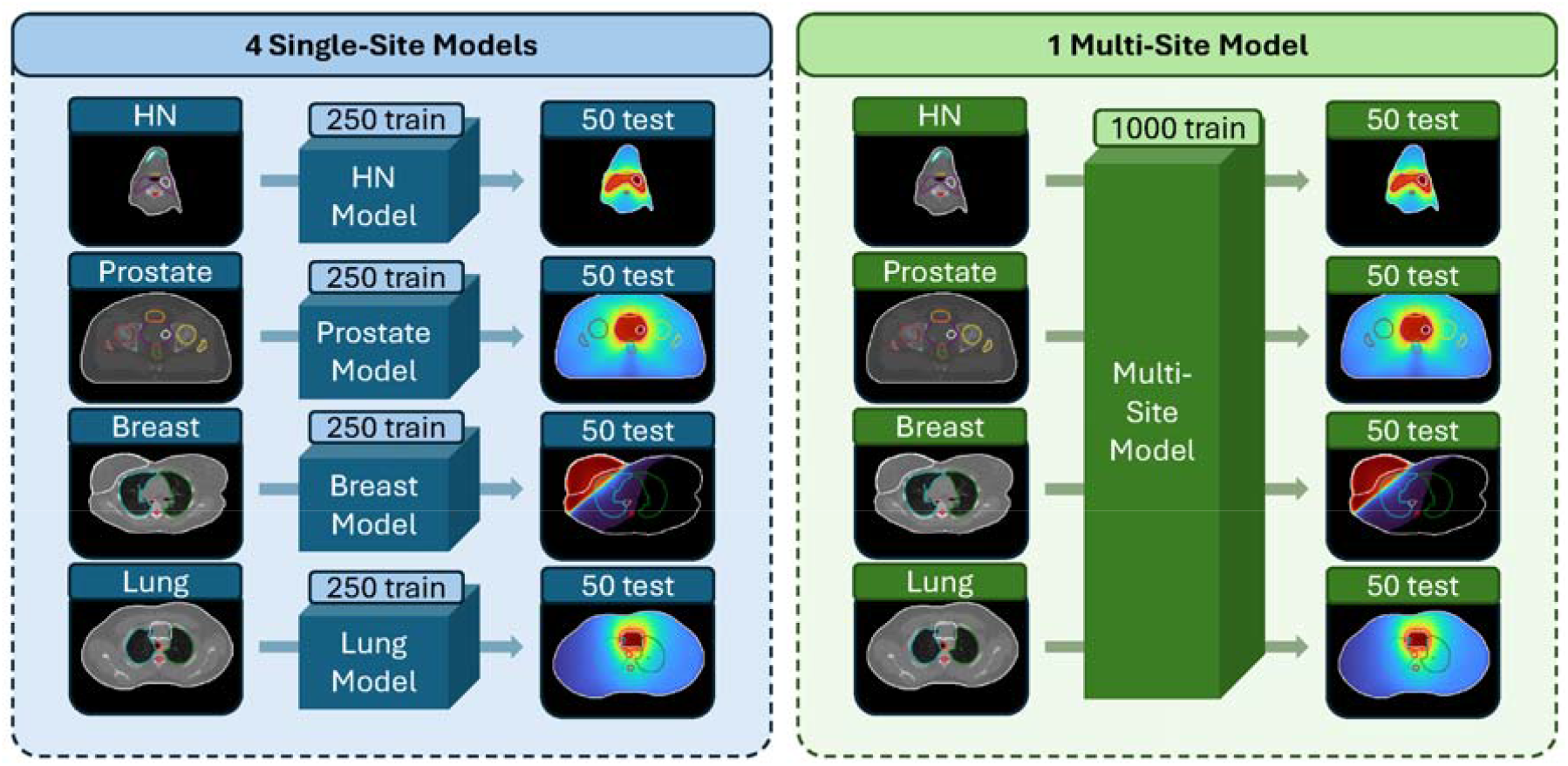
Schematic of the single-site models and multi-site models training. (Left) Single-site models, where four individual models were independently trained on 250 cases from each anatomical site (head and neck, prostate, breast, and lung); and (Right) Universal multi-site model, where a single model was trained on the consolidated dataset of 1,000 cases (250 cases per site). Both paradigms were evaluated on the same 200 seen site cases (50 per site) to ensure a direct comparison of performance. This experimental design serves to evaluate whether a single, beam-aware model can achieve clinical parity with specialized, site-specific models while significantly reducing the technical overhead of model management.

**Figure 4.**
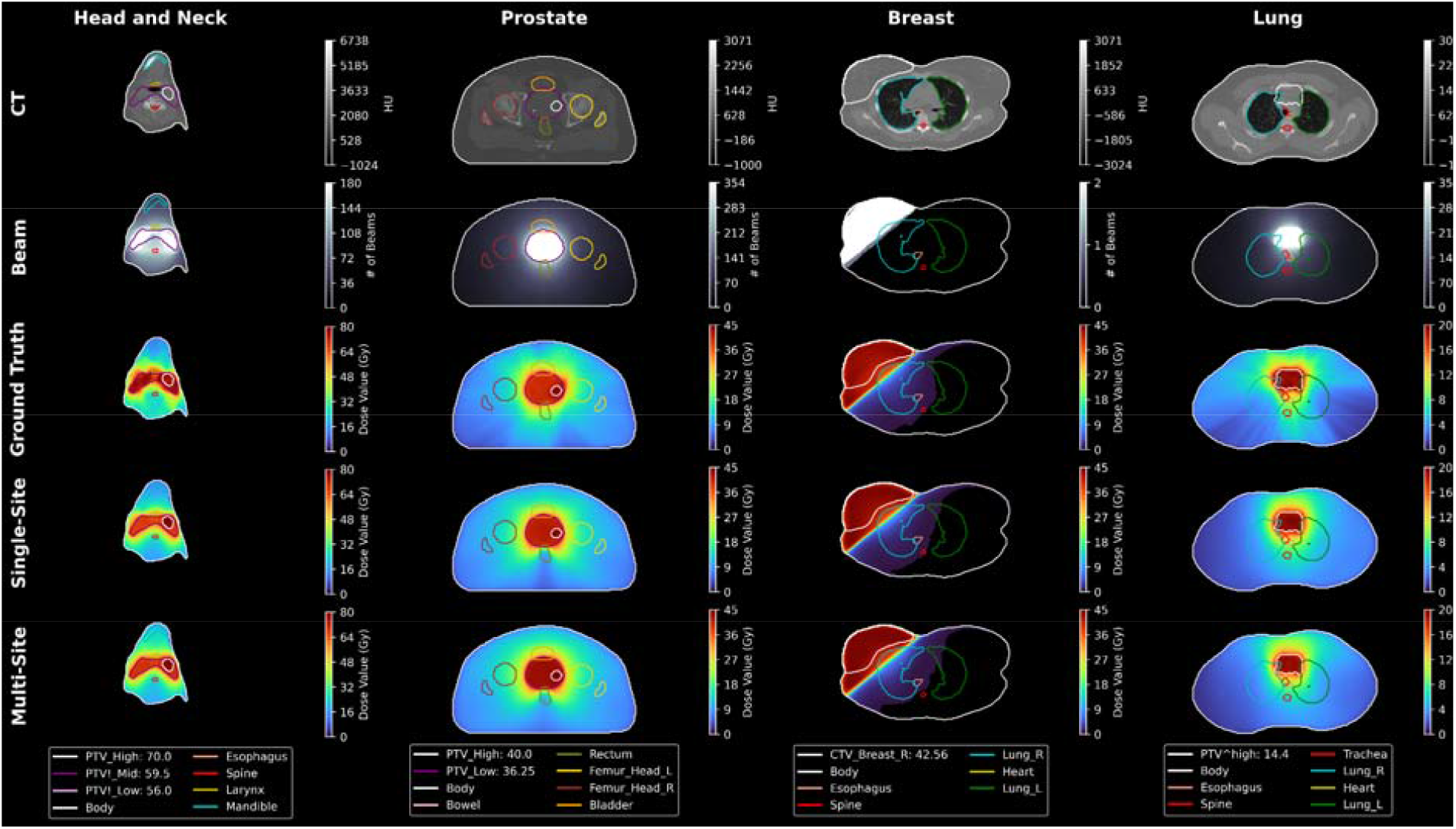
Representative dose prediction results across seen anatomical sites. Qualitative comparison of the input features and predicted dose distributions for the head and neck (HN), prostate, breast, and lung cohorts. For each site, the input CT, beam geometry map, and anatomical contours are displayed alongside the ground-truth dose and corresponding predictions from the site-specific and universal multi-site models. All results are shown for the prescription-labeled (Gy) models, with axial views representing typical cases from the 200-case independent testing set. The color bar indicates absolute dose values in Gy. The visual agreement between the predictions and ground-truth distributions demonstrates the framework’s ability to predict dose distribution across diverse anatomical regions and delivery geometries.

**Figure 4.**
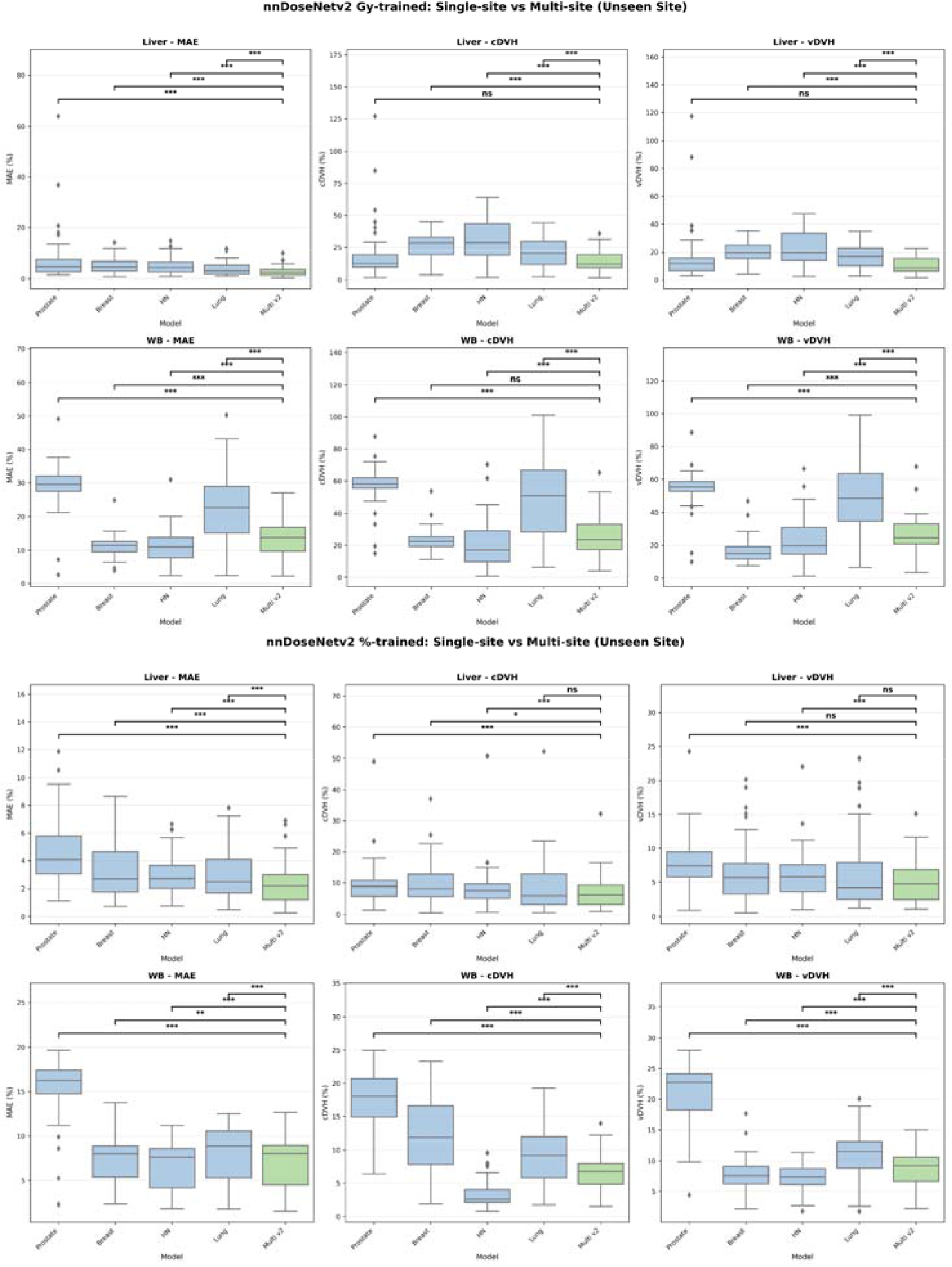
Generalizability of site-specific and universal multi-site models to unseen anatomical sites. Comparative performance of site-specific models (Blue) and the universal multi-site framework (Green) when validated on independent, unseen anatomical cohorts (Liver and Whole Brain). (Top two rows) Comparison of models utilizing the prescription-labeled (Gy) training format. (Bottom two rows) Comparison of models utilizing the percentage-labeled (%) training format. Performance is quantified via three distinct error-based metrics, where lower values indicate higher prediction accuracy: (Left) MAE, representing voxel-to-voxel dose differences. (Mid) cDVH representing difference in clinically relevant DVH metrics. (Right) vDVH representing difference in DVH curve. Statistical significance was determined using Wilcoxon signed-rank test (ns: not significant, * p < 0.05, ** p < 0.01, *** p < 0.001, and **** p < 0.0001.)

Consistent with the nnU-Net^32^ training framework, all nnDoseNet^5^ models were trained using 5-fold cross-validation, meaning that training occurred using 200 plans and cross-validated occurred on 50 plans for each fold. To ensure a fair and direct comparison between the single-site and multi-site training approaches, identical plans were in same fold during cross-validation. This guarantees that for any given fold, both the specialized and generalized models were trained on the exact same set of patient cases.

#### 2.3.2 Model Input

All models used the same 4-channel input which includes the CT, targets, OARs, and beam maps. All of the training configurations use the default nnDoseNet auto-configuration settings, which were optimized prior to this work. All models trained for this work use a 3D U-Net with depths of 6, batch size of 2 and stochastic gradient descent with momentum of 0.9. Patch sizes were determined for each model using the self-configuring process outlined in the nnU-net workflow.

#### 2.3.3 Loss Function

Default loss function of nnDoseNet^5^, the combination loss of MSE and DVH loss is utilized in this project. DVH loss, proposed by Wang et al.^26^, integrates the clinically relevant metrics, value-DVH (vDVH) and criteria-DVH (cDVH). The vDVH (equation 1) represents the dose gradient in contours while the cDVH (Equation 2) represents a specific dose value within a relevant volume of the contours. 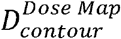represents the map of the ground truth (GT) or prediction (Pred) in certain contour (target or oar). N represents the number of contours and n represents the number of voxels within the contour. *Pr_x_* (D) represents the highest x percentile of dose value in the dose map, D.

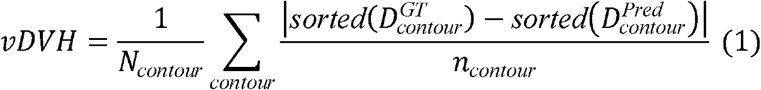

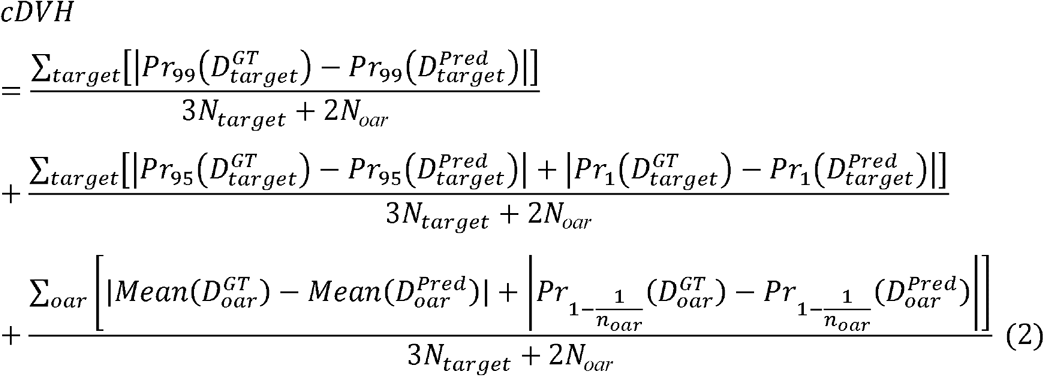

### 2.4 Post-processing and Evaluation

We compared the prediction of all 4×2 single-site models to the 1×2 multi-site models in both the Gy-value trained (Gy out) model and the percentage trained (percentage out) model. In addition to trained sites, our models were evaluated on the two unseen disease sites of liver and whole-brain (WB) cases, with each site represented by 50 cases.

Voxel-wise mean absolute error (MAE) and clinically relevant dose-volume histogram metrics (vDVH and cDVH) are utilized for evaluation. To facilitate comparison across multiple prescriptions, error metrics were normalized to the percentage of the prescribed dose; lower values denote higher prediction accuracy relative to the clinically delivered plans (ground truth). All tests of significance used paired Wilcoxon Signed-Rank Test when comparing the performance between two models.

### 2.5 The difference between nnDoseNet1 and nnDoseNet2

The core philosophy of the framework remains unchanged, to establish an automated pipeline for dose prediction using DICOM or NIfTI datasets. However, nnDoseNetv2 introduces significant upgrades to the data extraction module, most notably the ability to extract beam geometry directly from DICOM plan files. Computational efficiency was enhanced in both preprocessing and training by allowing the use of dose grid resolution rather than the typically higher CT resolution, and by cropping the input volume to exclude regions distant from the target. Finally, the input channel configuration was optimized; by leveraging the defined ROI, the beam map was integrated without increasing the input dimensionality, maintaining a four-channel architecture.

## 3 Results

### 3.1 Single-site vs multi-site model on seen site

The top and middle rows of Figure 5 present models trained using absolute dose (Gy) and percentage of prescription, respectively. In both configurations, the multi-site model (green) exhibits lower overall deviation. Furthermore, it shows no statistically significant difference compared to single-site models (p-value of MAE, vDVH and cDVH of prostate: 0.82, 0.89, 0.63; breast: 0.02, 0.87, 0.97; lung: 0.06, 0.30, 0.47), with the exception of HN cases (p < 0.01 for all metrics). For instance, the multi-site model achieved an MAE of 1.24 ± 0.49% for breast cases, which is comparable to the single-site breast model (1.29 ± 0.64%). Similarly, for HN cases, the multi-site model yielded an MAE of 1.09 ± 0.36%, closely matching the single-site result of 1.05 ± 0.36%. This indicates that the multi-site model achieves performance comparable to site-specific models while demonstrating greater stability.

**Figure 5.**
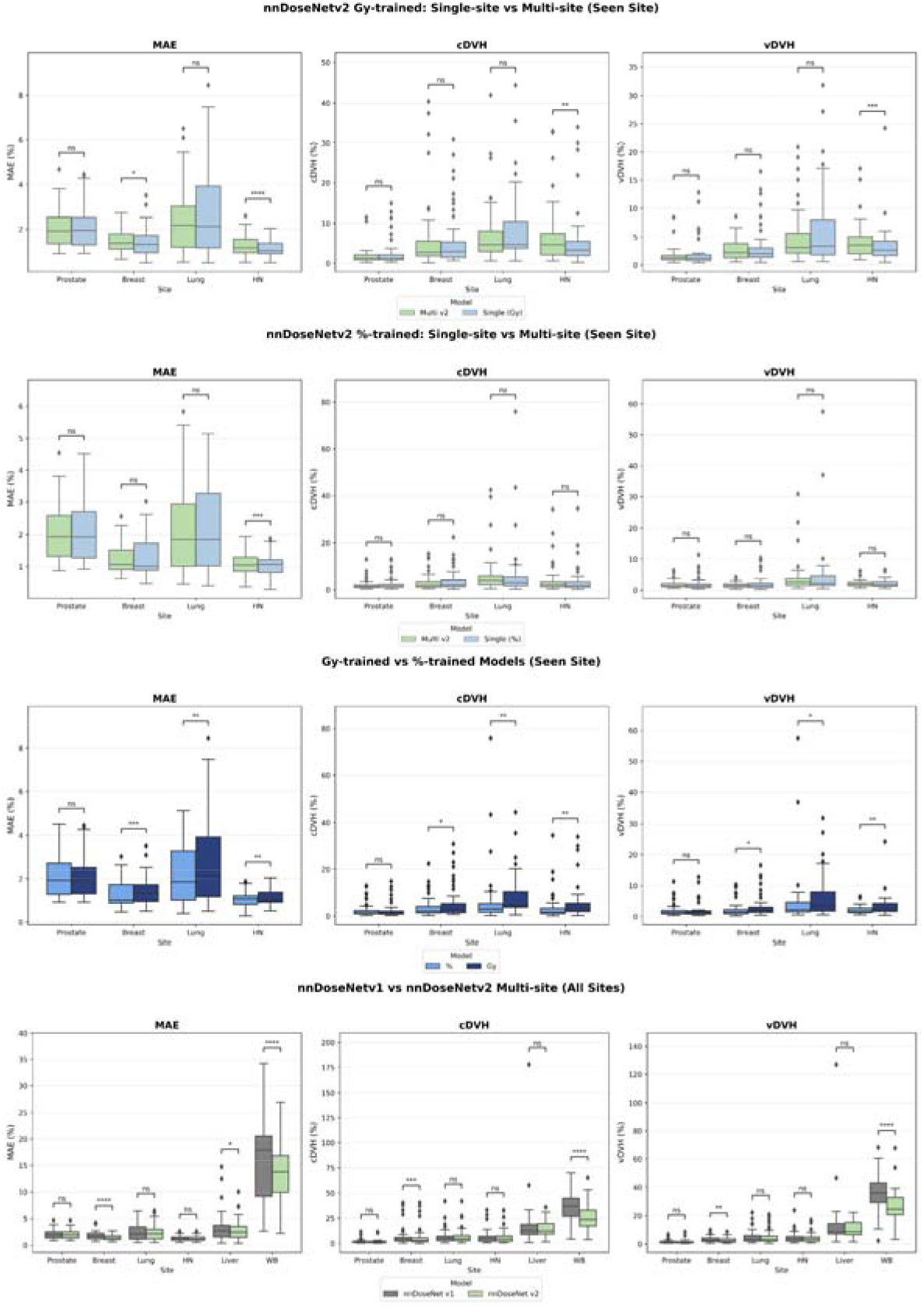
Evaluation of model scalability and beam-aware generalizability. (Rows 1–2) Site-specific vs. Multi-site performance: Comparison between specialized single-site models (Blue) and the universal multi-site model (Green) utilizing absolute dose (Gy; Row 1) and normalized dose (%; Row 2) training objectives. (Row 3) Labeling strategy comparison: Performance evaluation between Gy-labeled and percentage-labeled multi-site models across the four primary training sites. (Row 4) Impact of beam geometry on generalizability: Comparison between the baseline multi-site framework (nnDoseNet v1; Grey) and the upgraded beam-aware framework (nnDoseNet v2; Green). The integration of machine-specific beam geometry in nnDoseNet v2 results in significantly enhanced accuracy on anatomically unseen sites (Liver and Whole Brain) and the Breast cohort.Performance is quantified via three distinct error-based metrics, where lower values indicate higher prediction accuracy: (Left) MAE, representing voxel-to-voxel dose differences. (Mid) cDVH representing differece in clinically relevant DVH metrics. (Right) vDVH representing differece in DVH curve. Statistical significance was determined using Wilcoxon signed-rank test (ns: not significant, * p < 0.05, ** p < 0.01, *** p < 0.001, and **** p < 0.0001.)

When comparing the two multi-site approaches (Figure 5, bottom row), the percentage-trained model significantly outperforms the Gy-trained model across all trained sites (p-value of MAE, vDVH and cDVH of Breast: <0.001, 0.01, 0.01; Lung: <0.01, 0.01, <0.01; HN: <0.01, <0.01, <0.01), with the exception of prostate plans. For example, in Breast cases, the percentage-trained model achieved a VDVH error of 1.64 ± 0.92%, whereas the Gy-trained model resulted in a higher error of 2.71 ± 1.87%. However, for Prostate cases, the performance was nearly identical, with the percentage-trained model yielding an MAE of 2.07 ± 0.84% compared to 2.04 ± 0.83% for the Gy-trained model.

### 3.2 Multi-site model prediction on unseen sites

Two multi-site models were evaluated on two unseen anatomical sites, WB and Liver, with each test set consisting of 50 clinically delivered plans. For Liver cases (Figure 6), the multi-site model (MAE: 2.46 ± 1.49%) significantly outperformed most single-site models (p < 0.001), such as the Breast model (MAE: 3.47 ± 1.98%) and the Prostate model (MAE: 4.67 ± 2.33%). The only exceptions were the Gy-trained prostate model and the percentage-trained lung model, which showed no statistical significance (p > 0.05) difference compared to the multi-site model on DVH-based metrics.

Regarding WB patients, the multi-site models significantly (p < 0.001) outperformed the Prostate and Lung models. Specifically, the multi-site model achieved an MAE of 6.97 ± 3.04%, which was markedly superior to the Prostate model (15.41 ± 3.41%) and the Lung model (8.01 ± 3.14%). However, the single-site HN model achieved slightly better results (MAE: 6.69 ± 2.71%) than the multi-site model across most metrics.

### 3.3 nnDoseNetv1 vs nDoseNet2 on seen and unseen site

The primary distinction between nnDoseNetv1 and nnDoseNetv2 lies in the incorporation of beam geometry. This feature provides critical context regarding the treatment approach and spatial dose distribution. As shown in bottom row of Figure 5, nnDoseNetv2 significantly (p < 0.05) outperforms v1, particularly on unseen anatomical sites. For example, on the unseen WB dataset using Gy-trained models, nnDoseNetv2 reduced the MAE to 13.61 ± 4.98%, compared to 16.13 ± 7.33% for nnDoseNetv1.

Similarly, for the unseen Liver dataset, v2 achieved an MAE of 2.92 ± 1.95%, improving upon the v1 result of 3.29 ± 2.71%.

## 4 Discussion

In this study, we introduced a scalable training approach for radiotherapy dose prediction that utilizes beam geometry to bridge the gap between disparate anatomical sites and treatment approach. By utilizing an extensive cohort of 1,300 cases across six primary disease sites, we demonstrated that a single, universal model can maintain parity with specialized, site-specific models. To our knowledge, this is the first study to train a single deep learning model on a heterogeneous dataset encompassing a wide prescription range of 1.5 to 84 Gy, four distinct anatomical sites, and three different treatment modalities. Furthermore, we are the first to demonstrate the framework’s zero-shot generalizability by validating the model on out-of-distribution disease sites that were entirely excluded from the training phase.

A total of 1,000 clinically delivered RT plans across four distinct treatment sites were used to train our multi-site model, making it a single model trained with largest clinically delivered dataset for deep learning-based dose prediction reported to date. Our results show that the multi-site model performs comparably to the single-site models on the site seen, and even on liver cases, an unseen site that have similar anatomical structure to training sites. Even though single-site models trained with breast and HN dataset had a better performance than the multi-site model on WB site. This outcome is likely due to domain similarities: Breast and WB cases share comparable beam geometries, while HN cases share common OARs with WB cases. Although the multi-site model did not uniformly outperform the single-site models on their respective domains, it offers significant practical and clinical advantages. From an operational standpoint, a single, unified model is far easier to deploy and maintain than multiple specialized models. This approach streamlines the clinical prediction workflow and reduces the potential for user error, as clinicians can utilize the same tool regardless of the treatment site.

The heterogenous dataset training method effectively enlarges the training dataset by pooing the heterogenous data and further, it improves the generalizability and robustness of trained model. Furthermore, the multi-site training approach effectively increases the training data by pooling cases from different sites. This diversity acts as a natural regularizer, preventing the model from overfitting to site-specific nuances and enhancing its robustness. By training on a wide variety of anatomies, the model learns more generalizable features of dose-anatomy relationships and can implicitly share knowledge—such as strategies for sparing critical organs—across different anatomical contexts. Consequently, it is better equipped to handle atypical patient anatomy or rare clinical presentations. Perhaps the most significant advantage is the model’s potential for transfer learning; it can serve as a powerful foundation model that can be fine-tuned with a much smaller, site-specific dataset, drastically lowering the barrier to entry for adopting automated planning solutions in new clinics or for novel treatment types.

The incorporation of beam geometry improved the sharpness of the dose fall-off at the field edge, particularly for IMRT and 3D-CRT, which utilize more complex, conformal dose distributions. Initially, BEV was utilized in generating beam maps among all types of plans (IMRT, VMAT and 3D-CRT). But the predictions utilizing BEV fail on the breast cancer cases with low prescription doses and few beams (3D-CRT plans) where the dose should have cross-sectional shape more of field than conformed closely to the tumor volume. We also found that the WB cases performed better using the breast model and this is likely due to similarities in beam geometry between the two disease sites. These findings indicate that the model is highly reliant on beam geometry to produce a more robust and accurate dose distribution and the possibility to alter a predicted plan by inputting a different treatment geometry. For example, one could predict a VMAT dose distribution using the geometry from an IMRT plan, and vice versa. By incorporating machine-specific information into the input space, a single model can be effectively trained on heterogeneous datasets, marking a significant step toward global dose prediction.

Potential bias was found when training models only using Gy-labeled datasets and we observed that both single-site and multi-site perform better on prostate cases compared to other sites (Figure 5, top row). We suspect the reason might be that the model tends to prioritize the higher prescribed plan in training, which can efficiently lower the loss in the beginning. Therefore, we trained another set of models using percentage-labeled dataset, where we normalize the target mask and dose maps to percentile of prescription to eliminate the bias on higher prescribed plan. (Figure 5, bottom row) All other single-site models beside prostate, have significant (p < 0.05) better performance on cDVH and vDVH using percentage-trained model, that support our previous conjecture.

In terms of computational efficiency, we also significantly shortened the prediction time. By resampling all images to the dose resolution and cropping input channels around the target, the models can predict a full dose distribution within approximately 10 seconds on a 24 GB Titan RTX GPU. This is a substantial improvement over the 5-minute prediction time reported in our previous work and applies to both the single-site and multi-site models.

In addition to being trained on four primary disease sites, this study has several limitations. First, all data were derived from a single institutional planning environment, and the models may learn institution-specific planning preferences and contouring conventions rather than universally generalizable dose–anatomy relationships. Second, the clinically delivered dose was used as ground truth, which reflects historical practice and variability and does not necessarily represent an optimal reference plan. Third, while beam geometry was incorporated, the RTPLAN-derived beam map is a simplified surrogate that may not capture modulation and deliverability details (e.g., control-point fluence/MLC sequencing), which is important given the model’s reliance on the geometry channel. Finally, evaluation was based on MAE and DVH-derived metrics which quantify similarity to delivered dose but do not fully establish clinical acceptability, constraint satisfaction, or deliverability.

## 5 Conclusion

In summary, this study demonstrates the feasibility of training a single multi-site dose prediction model within the nnDoseNet framework using heterogeneous prescriptions and beam geometries, including an explicit beam-geometry input derived from DICOM plan files. Across the evaluated disease sites, the multi-site model achieved performance comparable to site-specific models while providing improved robustness in out-of-distribution testing, and the beam-geometry–enabled approach (nnDoseNetv2) improved prediction accuracy on unseen sites relative to the prior framework. Future work will focus on validation using multi-institutional datasets and expansion to additional disease sites to further assess generalizability.

## Data Availability

All data produced in the present study are available upon reasonable request to the authors

